# Waking to Brain Health: How Chronotype Relates to Brain Structure in Young Adults

**DOI:** 10.1101/2025.05.29.25328547

**Authors:** Iman Beheshti, Odelia Elkana

## Abstract

**Introduction:** Sleep patterns are critical for brain health and functioning, influencing mental health, learning, memory, and emotional regulation, particularly in young adults. However, the effects of chronotype, sleep quality, and daytime sleepiness on brain structure remain poorly understood. This study aimed to examine anatomical brain changes in individuals with early chronotype (EC) compared to those with late chronotype (LC). Early and late chronotypes, which refer to preferences for early or late sleep-wake times, represent distinct biological rhythms that may impact brain structure and function.

**Methods:** We used anatomical magnetic resonance imaging (MRI) to identify morphological and macroscopic brain differences between 68 young, healthy EC individuals and 68 age– and sex-matched LC participants. Two widely used atrophy estimation pipelines, voxel-based morphometry (VBM) and cortical thickness analysis, were employed to assess regional brain structure differences between the two groups.

**Results:** Whole-brain VBM analysis revealed decreased gray matter volume in LC individuals, specifically in the left cerebellum posterior lobe, left declive, and left cerebellum crus I. Additionally, linear regression models showed significant cortical thinning in the left caudal anterior cingulate, right caudal anterior cingulate, and right lateral occipital regions in the LC group compared to the EC group. However, there was no significant difference between the two groups in terms of white matter morphology or brain aging.

**Discussion:** Our findings suggest that chronotype-related differences in sleep patterns are associated with significant structural alterations in gray matter volume and cortical thickness. These changes may reflect the impact of chronic circadian misalignment on neurobiological integrity, even in young, healthy individuals. Findings emphasize the importance of considering sleep timing as a critical factor in brain health and lay the groundwork for future longitudinal studies.

## 1. Introduction

Chronotype refers to individual differences in preferred sleep–wake timing, reflecting an individual’s endogenous circadian rhythm. Chronotypes are commonly categorized as early chronotypes (EC) or “morning types,” and late chronotypes (LC) or “evening types.” EC individuals tend to wake and function optimally in the morning, whereas LC individuals show peak alertness and cognitive performance later in the day[1]. These preferences are largely influenced by genetic factors (e.g., *PER3*) and are associated with key physiological processes, including hormonal secretion, metabolism, cognitive efficiency, and affective regulation [2]. While relatively stable, chronotype shifts across the lifespan, toward eveningness during adolescence and toward morningness with aging. Gender differences have also been observed, with females more likely to prefer earlier schedules.

In contemporary environments, social and behavioral demands, such as artificial lighting, technology use, and rigid work schedules, can exacerbate misalignment between one’s biological rhythm and external constraints. This circadian misalignment, often described as social jetlag, is particularly pronounced in late chronotypes and has been linked to increased sleep debt, greater emotional reactivity, impaired cognitive performance, and dysregulation of neuroendocrine and immune function (e.g., elevated evening cortisol, altered melatonin secretion, heightened inflammation) [3, 4].

Despite growing evidence linking circadian rhythm and sleep quality to brain function, the structural neural correlates of chronotype remain underexplored—especially in healthy young adults, who may be uniquely vulnerable to chronic sleep–wake misalignment. Emerging neuroimaging studies have revealed chronotype-related differences in gray matter volume and functional connectivity, particularly in regions such as the precuneus, insula, thalamus, orbitofrontal cortex, and lateral occipital cortex [5-7]. However, findings have been inconsistent, and few studies have combined both volumetric and cortical thickness methods or controlled for demographic and circadian variables [8].

Of particular interest is the anterior cingulate cortex (ACC), a region centrally involved in emotional regulation and cognitive control. While functional differences in the ACC have been reported across chronotypes, few studies have investigated structural alterations such as cortical thinning in this region. Similarly, although the occipital cortex is classically associated with visual processing, recent work has linked it to circadian light sensitivity, melatonin suppression, and functional connectivity shifts in evening types, suggesting that occipital morphology may also vary by chronotype [9-11]. In addition, the cerebellum, long viewed as a motor-related structure, has gained recognition for its role in higher-order cognitive and affective functions, including attention, timing, and emotional modulation [12, 13]. Yet, this region remains largely overlooked in chronotype research.

Beyond regional brain morphology, a novel approach to understanding individual variability involves brain age estimation models, which use machine learning to derive a neurobiological age based on MRI data. Deviations between predicted and chronological age can serve as early markers of accelerated brain aging or diminished brain maintenance [14, 15]. Although sleep quality has been associated with brain aging, the potential contribution of chronotype and sleep timing to brain aging trajectories remains largely unexplored

### Present Study

To address these gaps, the present study investigated neuroanatomical differences between early and late chronotypes in a sample of healthy young adults. High-resolution structural MRI was used to assess brain morphology via two complementary techniques: voxel-based morphometry (VBM) and cortical thickness analysis, enabling a detailed evaluation of gray matter volume and cortical integrity across the brain. In addition, we applied a brain age estimation model to examine whether LC individuals exhibit signs of accelerated brain aging.

Our key objectives were to:

1. Determine whether EC and LC individuals differ in regional gray matter volume and cortical thickness.
2. Assess whether LC individuals show greater deviation between estimated and chronological brain age.
3. Explore whether sleep-related variables (e.g., sleep quality, duration, circadian alignment) are associated with individual differences in brain structure.

Based on prior research and emerging neurobiological models, we hypothesized that:

1. LC individuals would exhibit reduced gray matter volume and cortical thinning, particularly in regions implicated in emotional regulation and sensory, circadian integration, such as the ACC, lateral occipital cortex, and posterior cerebellar lobules.
2. LC individuals would show greater brain age deviation compared to EC individuals, suggesting possible early signs of accelerated brain aging.
3. Sleep-related variables, including sleep quality, duration, and alignment, would be associated with structural differences in brain morphology, particularly in regions showing group-level chronotype effects.

To address these objectives, we conducted a structural MRI study comparing EC and LC individuals, using a multimodal neuroimaging approach. The following section describes the participant selection criteria, MRI acquisition protocol, preprocessing steps, brain age estimation framework, and statistical analysis procedures.

## 2. Materials and Methods

### 2.1. Participants and MRI Acquisition

We recruited 68 young, healthy individuals with an EC and an equal number of age– and sex-matched participants with a LC from the OpenNeuro dataset (https://openneuro.org/datasets/ds003826/versions/1.1.0/3.0.0; accessed on March 1, 2025). All participants were right-handed, had normal or corrected-to-normal vision, and had no history of neurological or psychiatric disorders. None were taking medications at the time of the study. The participants were between 19 and 35 years of age, had no prior history of shift work, and had not undertaken flights crossing more than two time zones in the preceding two months. Additional inclusion criteria included the absence of excessive daytime sleepiness, assessed using the Epworth Sleepiness Scale (ESS), with a threshold score of ≤10, indicating normal daytime alertness. The ESS is a standardized tool that evaluates the likelihood of dozing off in various daily situations. Lower ESS scores indicate a state of wakefulness and low susceptibility to daytime sleep, whereas higher scores suggest an increased likelihood of daytime drowsiness, potentially indicative of underlying sleep disorders or insufficient sleep. Sleep quality was assessed using the Pittsburgh Sleep Quality Index (PSQI), with a cutoff score of ≤5, reflecting good subjective sleep quality. The PSQI is a widely utilized instrument that evaluates multiple dimensions of sleep, including duration, disturbances, latency, and overall satisfaction. Higher PSQI scores are indicative of poorer sleep quality. Participants were required to maintain a regular sleep schedule without evidence of sleep deprivation, defined as obtaining 6 to 9 hours of sleep per night.

Chronotype classification was conducted using the Chronotype Questionnaire (ChQ), which includes multiple scales to assess circadian preferences [16, 17]. The morningness-eveningness scale (ChQ-ME) quantifies an individual’s preference for morning versus evening activities, with scores ranging from 16 to 86. Additionally, the amplitude scale (ChQ-AM), a component of the Chronotype Questionnaire, was used to measure the strength and variability of an individual’s circadian rhythm [16, 17]. A higher ChQ-ME subscale score indicates a stronger evening preference, while a higher ChQ-AM score signifies greater variability in mental processes throughout the day.

In this study, participants were classified as morning or evening types based on their ChQ-ME scores:

- **EC** (early chronotype), with ChQ-ME scores between 11 and 21.
- **LC** (late chronotype), with ChQ-ME scores between 22 and 32.

The threshold of 21 points for classifying early and late chronotypes was determined based on the distribution of morningness-eveningness scores [18]. This distinct separation in the data rendered the creation of an intermediate group unnecessary, as the two-group classification effectively captured the clear distinction between preferences for morning and evening activities.

Anatomical MRI scans were acquired using a 3T Siemens Skyra scanner with a GR_IR sequence (repetition time = 2.3 s, echo time = 2.98 ms, inversion time = 0.9 s, flip angle = 9°, slice thickness = 1.1 mm, base resolution = 256, matrix = 248 × 256, field of view = 24 cm × 24 cm, acquisition matrix = 248, reconstruction matrix = 256, number of slices = 180, partial Fourier = 1, parallel reduction factor = 2, and scan duration = 3.19 minutes). Further details regarding demographic and clinical assessments, as well as MRI acquisition protocols, are provided in [18].

### 2.2. MRI Preprocessing

We processed the anatomical MRI data using the CAT12 toolbox (http://www.neuro.uni-jena.de/cat/, accessed on 20 February 2025), which extends the functionality of the Statistical Parametric Mapping (SPM15) software package (https://www.fil.ion.ucl.ac.uk/spm/software/spm25/, accessed on 20 February 2025). The preprocessing steps included bias field correction, segmentation of the MRI images into gray matter (GM), white matter (WM), and cerebrospinal fluid (CSF), DARTEL normalization to MNI space (voxel size = 1 mm × 1 mm × 1 mm), and modulation. We assessed image quality through visual inspection and used the “Check Homogeneity” feature in CAT12 to identify potential artifacts or variability in voxel intensity that might affect the accuracy of subsequent analyses. We applied an 8 mm full-width-half-maximum Gaussian kernel for spatial smoothing of the images. A detailed description of the VBM analysis methodology is provided elsewhere [19]. We also calculated the total intracranial volume (TIV) using CAT12. For VBM analysis, we used GM and WM density images. We obtained cortical thickness (CT) measurements using the CAT12 toolbox in combination with the Desikan-Killiany-Tourville (DKT) atlas, which defines 34 predefined regions of interest (ROIs) in each hemisphere to assess cortical thickness [20].

### 2.3 Brain age estimation framework

To estimate brain age for both the LC and EC groups, we used a pre-trained brain age prediction model developed from three datasets of healthy individuals: the IXI dataset (563 participants), OASIS (313 HC and 120 Alzheimer’s patients), and PPMI (373 Parkinson’s patients and 198 HC) [21]. A total of 1,054 healthy control (HC) T1-weighted MRI scans were used, which were divided into a training set (90% of participants, N=949) and a validation set (10% of participants, N=105). The model employed support vector regression (SVR) with a linear kernel, utilizing brain structural data such as GM, WM, CSF volumes, and other factors like scanner type, magnetic field strength, and sex as inputs. Chronological age was the target output. The model performed well on both the training set (MAE = 4.72 years, RMSE = 6.07 years) and the validation set (MAE = 4.63 years, RMSE = 5.88 years), indicating strong predictive accuracy. The final model, trained on the full dataset, was subsequently tested on the independent LC and EC datasets. The brain age prediction model typically yield an outcome referred to as the Brain-Predicted Age Difference (Brain-PAD), defined as the difference between an individual’s predicted brain age and their chronological age [14]. A positive Brain-PAD score reflects accelerated brain aging, whereas a negative score may indicate preserved or delayed brain aging relative to chronological age. This metric serves as an indicator of overall brain health [15].

### 2.4. Statistical Analysis

To assess structural differences between the EC and LC groups, independent t-tests were conducted on the GM and WM images using SPM25. A peak-level significance threshold of p < 0.001 (uncorrected) was applied, with clusters considered significant if they surpassed the false discovery rate (FDR)-corrected cluster-level threshold of q < 0.05. Voxel-based analysis was performed across the entire brain to identify regional variations in GM and WM. Additionally, changes in GM and WM were examined in relation to the ChQ-ME, ChQ-AM, ESS, and PSQI scores through multiple regression analysis in SPM25, employing an uncorrected threshold of p < 0.001. Age, sex, and TIV were included as covariates in the design matrix of the VBM analysis. Furthermore, to compare cortical thickness between the EC and LC groups, linear regression models were applied, controlling for age and sex as follows:

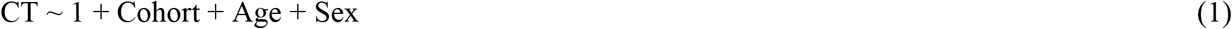

In this model, CT represents cortical thickness values in each region, and Cohort is the variable of interest that differentiates between the LC and EC groups. A total of 68 linear regression models were conducted.

## 3. Results

### 3.1 Clinical and demographical characteristics

No significant differences were found between the two groups in terms of age, sex distribution, ChQ-AM, PSQI, or ESS scores (p > 0.05 for all). However, a significant difference was observed in ChQ-ME scores (p < 0.0001), reflecting a pronounced divergence in morningness-eveningness preference, with the EC group reporting significantly higher morningness scores compared to the LC group. This suggests that the two groups exhibit distinct chronotypic preferences. Table 1 outlines the demographics and characteristics of EC individuals and 68 matched LC participants.

**Table 1:** Demographic Information of 68 Young, Healthy EC and 68 Age– and Sex-Matched LC Participants in the Study.

|  | EC | LC | p-value |
| --- | --- | --- | --- |
| Number | 68; 33.8% male | 68; 38.2% male | 0.72 |
| Age, years | $24.80 \pm 3.90$ | $23.85 \pm 3.51$ | 0.12 |
| ChQ-ME | $16.76 \pm 2.69$ | $26.67 \pm 2.61$ | $<0.0001$ |
| ChQ-AM | $20.42 \pm 3.97$ | $21.19 \pm 3.49$ | 0.23 |
| PSQI | $2.94 \pm 1.26$ | $3.21 \pm 1.52$ | 0.27 |
| ESS | $6.79 \pm 3.16$ | $7.33 \pm 3.83$ | 0.36 |

**Note:** EC = early chronotype; LC = late chronotype; ChQ-ME = morningness-eveningness scale of the Chronotype Questionnaire; ChQ-AM = amplitude scale of the Chronotype Questionnaire; PSQI = Pittsburgh Sleep Quality Index; ESS = Epworth Sleepiness Scale. For continuous variables, the t-test was used to assess differences between groups, while the $\chi^2$ test was used for categorical variables. There were no significant differences between EC and LC participants for age, ChQ-AM, PSQI, or ESS scores. The significant difference observed in ChQ-ME scores ( $p < 0.0001$ ) indicates a marked contrast in morningness-eveningness preference between the two groups.

### 3.2. Voxel-Based Morphometry Analysis

The VBM analysis revealed a significant decrease in GM in the LC group compared to the EC group. The most prominent region of GM reduction was identified in the left cerebellum, specifically within the cerebellum posterior lobe and the declive. This cluster, consisting of 4097 voxels, also extended into regions such as the cerebellum crus1, occipital lobe, and fusiform gyrus (Table 2 and Fig 1). No significant differences were observed in WM volume between the EC and LC groups (p < 0.001, uncorrected). Additionally, multiple regression analysis in SPM25 was conducted to assess the relationship between GM/WM changes and the ChQ-ME, ChQ-AM, ESS, and PSQI scores. No significant interactions were found between GM and WM volumes and these variables.

**Table 2:** Gray Matter Atrophy Cluster Identified by VBM Analysis (SPM25) in LC Participants Compared to Age– and Sex-Matched EC Group.

| Region | Cluster Size<br>(No. of Voxels) | q<br>(FDR) | Hemisphere | MNI<br>Coordinates<br>(x, y, z) | T Value<br>(Peak Voxel) |
| --- | --- | --- | --- | --- | --- |
| Cerebellum/Cerebellum Posterior<br>Lobe/Declive/Cerebellum_6/Cerebellum<br>Crus1 | 4097 | 0.005 | Left | -22, -69, -24 | 4.05 |

**Note:** MNI = Montreal Neurological Institute; FDR = false discovery rate.

**Figure 1.**
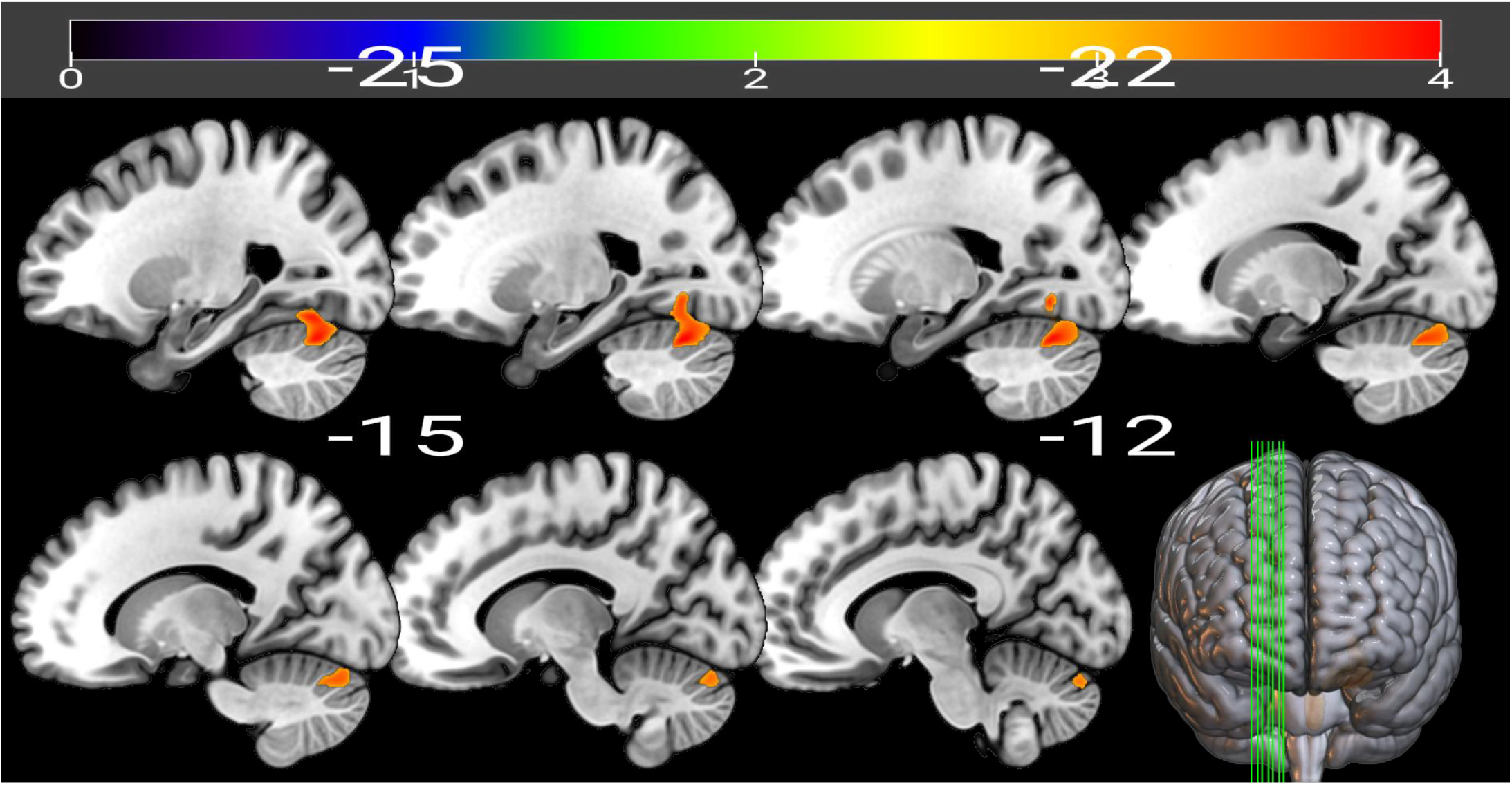
Illustration of Gray Matter Volume Reduction in LC Participants Compared to Age-and Sex-Matched EC Group via VBM in SPM25. Significant gray matter atrophy was observed in the left cerebellum, particularly within the cerebellum posterior lobe and declive. Regions with pronounced atrophy are shown in color. The T-map was created with an FDR-uncorrected threshold of p < 0.0001 and an extent threshold of k > 500. The color bar indicates the t-values for the comparison between the two groups.

### 3.2. Cortical thicknerss analysis

The linear regression analysis revealed significant differences in cortical thickness between the EC and LC groups. Specifically, the LC group showed decreased cortical thickness compared to the EC group in the left and right caudal anterior cingulate, and right lateral occipital regions. In contrast, the EC group had thinner cortical thickness than the LC group in the left temporal pole (Table 3 and Fig 2). Notable findings include significant reductions in the left caudal anterior cingulate (β = –0.106, t = –2.946, p = 0.0037), right caudal anterior cingulate (β = –0.100, t = –2.893, p = 0.0044), and right lateral occipital region (β = –0.035, t = –2.078, p = 0.039) for the EC group. Conversely, the EC group showed increased cortical thickness in the left temporal pole (β = 0.105, t = 2.020, p = 0.045). Age was a significant predictor of cortical thickness in three regions, with older age correlating to thinner cortical thickness, while sex influenced only the right caudal anterior cingulate. Figure 3 presents boxplots illustrating cortical thickness in regions where statistically significant differences were observed between the EC and LC groups, as determined by linear regression analysis.

**Table 3:** Regression Analysis Results for Cortical Thickness, Showing Significant Differences Between EC and LC Groups (p-value < 0.05 for Group).

| Model |  | left caudal<br>anterior cingulate | right caudal anterior<br>cingulate | right lateral<br>occipital regions | left temporal pole |
| --- | --- | --- | --- | --- | --- |
| Group | $\beta$ | -0.106 | -0.100 | -0.035 | 0.105 |
|  | t | -2.946 | -2.893 | -2.078 | 2.020 |
|  | p | 0.0037 | 0.0044 | 0.039 | 0.045 |
| Age | $\beta$ | -0.012 | -0.013 | -0.007 | 0.004 |
|  | t | -2.514 | -2.851 | -3.187 | 0.608 |
|  | p | 0.013 | 0.005 | 0.001 | 0.544 |
| Sex | $\beta$ | -0.019 | 0.073 | -0.002 | 0.013 |
|  | t | -0.521 | 2.063 | -0.161 | 0.251 |
|  | p | 0.602 | 0.0410 | 0.871 | 0.801 |

**Table 3:** Summary of Significant Regression Results for Cortical Thickness Regions and Their Associations with ChQ-ME, ChQ-AM, PSQI, and ESS.

| Outcome | Region | $\beta$ | $t$ | $p$ | $\beta$ | $t$ | $p$ | $\beta$ | $t$ | $p$ |
| --- | --- | --- | --- | --- | --- | --- | --- | --- | --- | --- |
| Variable |  | (Region) | (Region) | (Region) | (Age) | (Age) | (Age) | (Sex) | (Sex) | (Sex) |
| ChQ-ME | Right Caudal Anterior Cingulate Cortex | -7.584 | -3.321 | 0.001 | -0.286 | -2.235 | 0.027 | 0.858 | 0.871 | 0.386 |
| **ChQ-ME** | Left Temporal Pole | 4.172 | 2.671 | 0.009 | -0.208 | -1.641 | 0.103 | 0.225 | 0.229 | 0.819 |
| **ChQ-ME** | Left Caudal Anterior Cingulate Cortex | -5.620 | -2.516 | 0.013 | -0.256 | -1.982 | 0.050 | 0.180 | 0.182 | 0.856 |
| **ChQ-ME** | Right Lateral Occipital Cortex | -9.782 | -2.034 | 0.044 | -0.264 | -1.995 | 0.048 | 0.249 | 0.251 | 0.802 |
| **ChQ-AM** | Right Insula | -4.811 | -2.380 | 0.019 | -0.128 | -1.470 | 0.144 | 1.285 | 1.969 | 0.051 |
| **ChQ-AM** | Left Lateral Occipital Cortex | -7.296 | -2.235 | 0.027 | -0.102 | -1.200 | 0.232 | 1.448 | 2.192 | 0.030 |
| **ChQ-AM** | Left Caudal Anterior Cingulate Cortex | -3.161 | -2.129 | 0.035 | -0.110 | -1.275 | 0.205 | 1.185 | 1.810 | 0.073 |
| **PSQI** | Right Fusiform gyrus | -2.198 | -2.096 | 0.038 | -0.024 | -0.729 | 0.467 | 0.198 | 0.789 | 0.432 |
| **ESS** | Left Pars Orbitalis | -4.702 | -2.195 | 0.030 | -0.072 | -0.887 | 0.377 | 0.820 | 1.324 | 0.188 |

**Note:** ChQ-ME = morningness-eveningness scale of the Chronotype Questionnaire; ChQ-AM = amplitude scale of the Chronotype Questionnaire; PSQI = Pittsburgh Sleep Quality Index; ESS = Epworth Sleepiness Scale. The regression analyses were performed, where the dependent variables included ChQ-ME, ChQ-AM, PSQI, and ESS. Independent variables included age, sex, and cortical thickness measurements from various brain regions. Only the regions with p-values < 0.05 were considered statistically significant.

**Figure 2.**
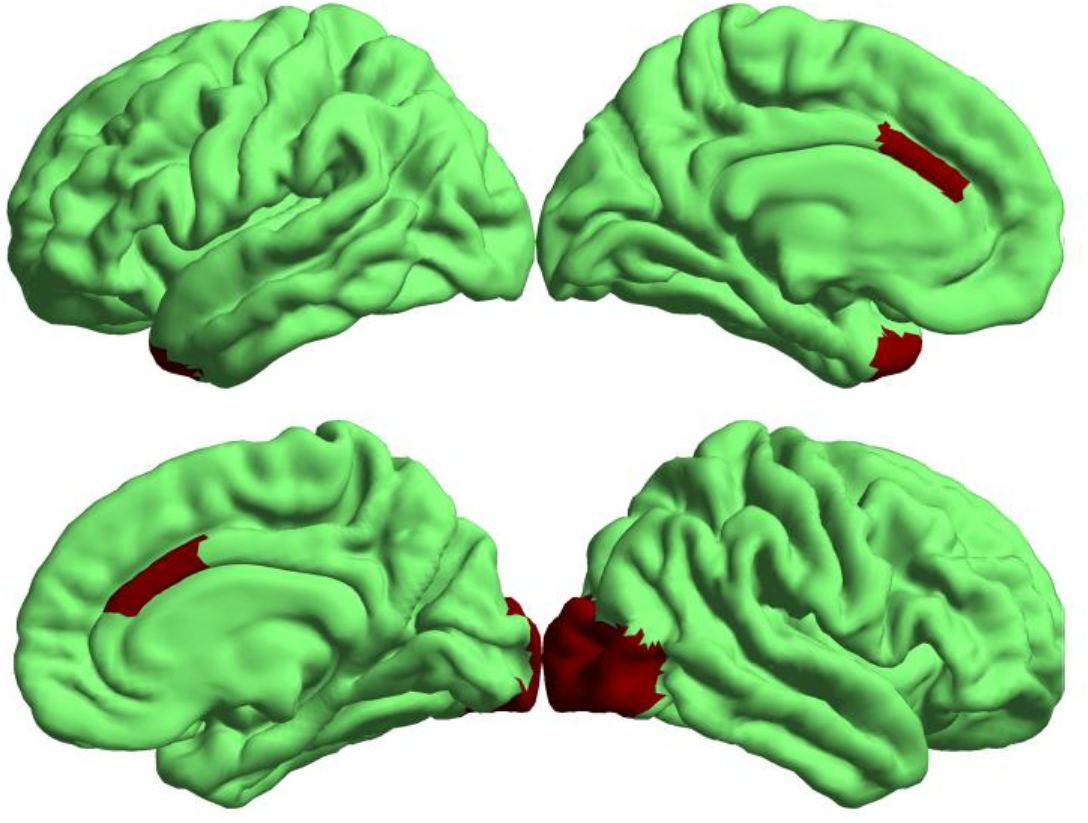
Visualization of significant cortical-thickness differences between EC and LC groups, derived from regression analysis adjusted for age and sex.

**Figure 3.**
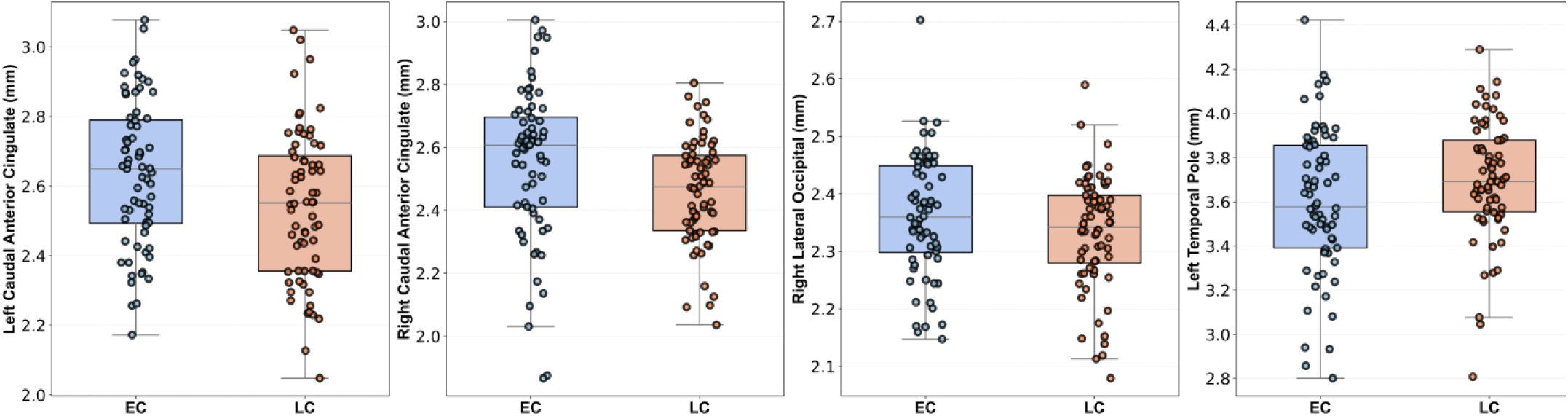
Boxplots of cortical thickness across different regions demonstrate statistically significant differences (p < 0.05) between the EC and LC groups, as determined by linear regression analysis, with age and sex as covariates.

**Figure 4.**
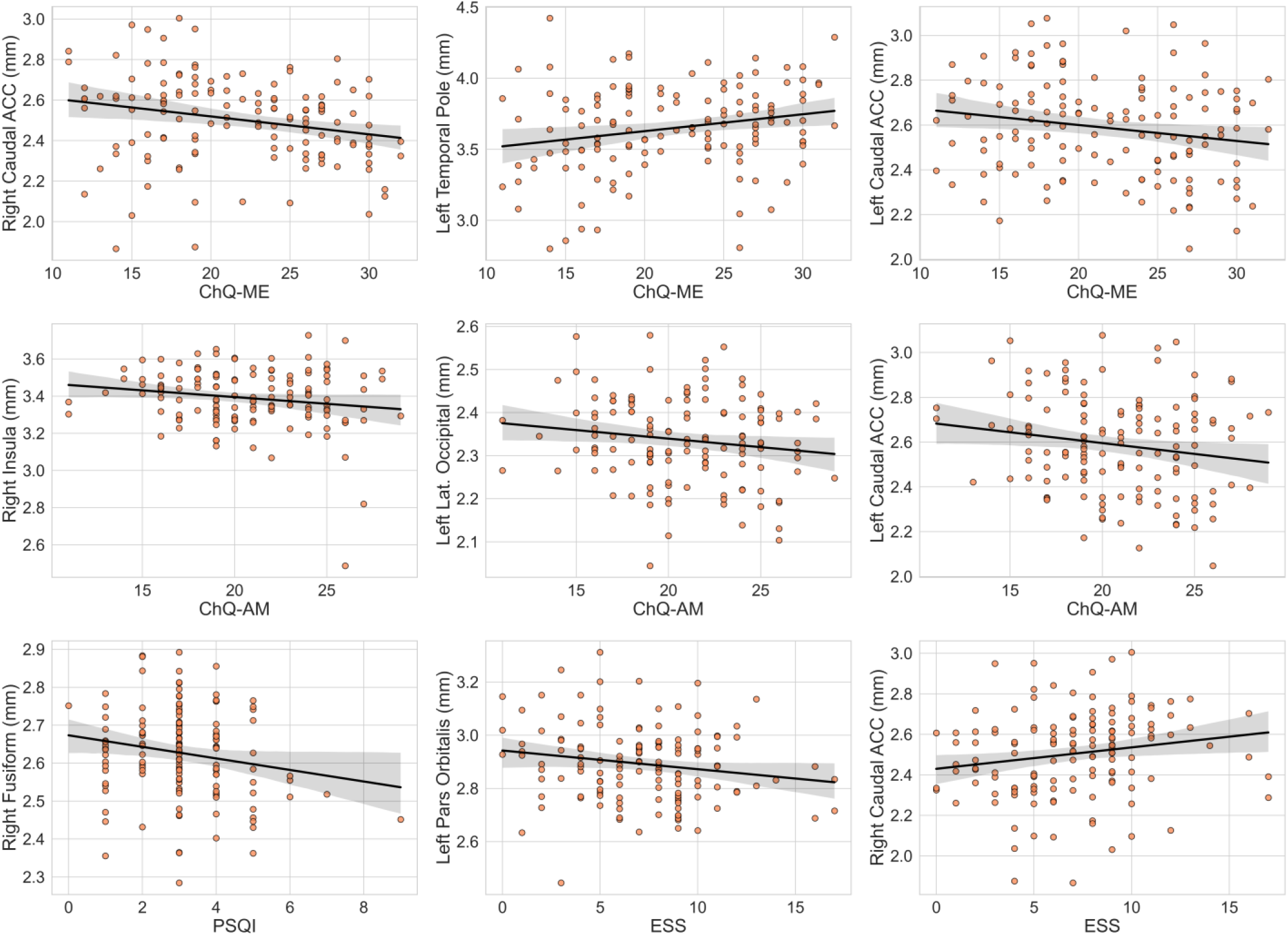
Scatter plots with regression lines for cortical thickness values that showed statistically significant associations with clinical variables (ChQ-ME, ChQ-AM, PSQI, and ESS). Statistical significance was determined using linear regression models between each clinical variable and cortical thickness across 68 brain regions. Age and sex were included as covariates in all regression models.

Regression analyses revealed a regionally specific relationship between CT measurements and multiple sleep-related dimensions. In analyses with ChQ-ME as the outcome, reduced CT in the right caudal anterior cingulate cortex was significantly associated with lower ChQ-ME scores (β = –7.584, t = –3.321, p = 0.001), paralleled by a similar negative association in the left caudal anterior cingulate cortex (β = –5.620, t = – 2.516, p = 0.013). In contrast, greater CT in the left temporal pole corresponded to higher ChQ-ME scores (β = 4.172, t = 2.671, p = 0.009). Additionally, the right lateral occipital cortex exhibited a negative association with ChQ-ME (β = –9.782, t = –2.034, p = 0.044). For the amplitude dimension of the ChQ-AM, decreased CT in the right insula (β = –4.811, t = –2.380, p = 0.019), left lateral occipital cortex (β = – 7.296, t = –2.235, p = 0.027), and left caudal anterior cingulate cortex (β = –3.161, t = –2.129, p = 0.035) were each significantly associated with lower amplitude scores. Sleep quality and daytime sleepiness also demonstrated cortical correlates. Specifically, diminished CT in the right fusiform gyrus was linked to poorer sleep quality as measured by the PSQI (β = –2.198, t = –2.096, p = 0.038), whereas reduced CT in the left pars orbitalis was associated with higher scores on the ESS (β = –4.702, t = –2.195, p = 0.030). All models included age and sex as covariates, with age showing modest effects in some regions, while sex did not emerge as a significant predictor.

### 3.3 Brain Age Estimation

No significant differences in brain-predicted age difference (Brain-PAD) were found between the LC (M =

0.6 ± 4.3 years) and EC (M = 0.4 ± 5.2 years) groups (t (134) = –1.45, p = 0.15). These findings suggest that, on average, the extent of deviation between predicted and chronological brain age does not differ significantly by chronotype, despite observable structural differences in gray matter volume and cortical thickness across regions.

## 4. Discussion

This study investigated the neuroanatomical correlates of chronotype in healthy young adults using voxel-based morphometry (VBM), cortical thickness analysis, and brain age estimation. The findings demonstrate that individuals with an LC exhibit specific regional alterations in brain structure compared to EC individuals, despite no differences in subjective sleep quality, daytime sleepiness, or WM volume.

Most notably, LC individuals exhibited reduced gray matter volume in the left cerebellum, including the posterior lobe, declive, and Crus I. Beyond its classical role in motor coordination, the cerebellum, particularly posterior regions such as Crus I, has been increasingly implicated in higher-order functions, including executive control, working memory, and emotion regulation. These functions are known to be sensitive to sleep timing and circadian integrity. Therefore, the structural reduction observed in LC individuals may reflect neuroplastic adaptations or vulnerabilities associated with chronic circadian misalignment, such as irregular sleep-wake cycles and persistent desynchronization between internal biological rhythms and external environmental demands.

Cortical thickness analyses further revealed bilateral thinning in the ACC, a region centrally involved in conflict monitoring, emotional awareness, and cognitive control [22, 23]. The structural vulnerability of the ACC among LC individuals is consistent with evidence suggesting reduced fronto-limbic connectivity in evening types, and may reflect diminished regulatory capacity even in the absence of clinical symptomatology [22, 23]. The finding of right lateral occipital thinning adds to emerging evidence that posterior cortical regions may be sensitive to circadian influences, potentially due to their involvement in visual processing and responsiveness to light cues that modulate circadian entrainment and melatonin suppression [9-11].

In contrast, brain age prediction did not reveal significant differences between LC and EC participants. This suggests that, at the global level of whole-brain structure, both chronotype groups exhibit comparable aging trajectories in young adulthood. Nevertheless, the regional gray matter and cortical thinning observed in LC individuals may represent early-stage structural variations that precede more widespread age-related changes. These findings underscore the importance of evaluating both global and region-specific markers when examining brain aging processes, especially in relation to circadian preference.

Interestingly, structural alterations in the LC group were present despite no significant group-level differences in PSQI or ESS scores. This implies that brain changes may occur independently of perceived sleep problems, and may instead reflect a stable, biologically driven trait. Alternatively, it is possible that conventional self-report measures lack the sensitivity to detect subtle dysfunctions in young, high-functioning individuals.

Our findings contribute both confirmatory and novel insights to the literature. While prior functional studies have implicated the ACC and cerebellum in chronotype-related variation, structural findings in these regions have been scarce [13]. The present study provides anatomical evidence supporting the role of the ACC and posterior cerebellum, particularly Crus I, in chronotype-sensitive brain organization. These regions may form part of a broader fronto-cerebellar regulatory network sensitive to the temporal dynamics of sleep and wakefulness. The lack of structural differences in prefrontal regions such as the dorsolateral PFC (DLPFC) may reflect developmental factors. Prefrontal myelination continues into the third decade of life, and structural divergences in these regions may only emerge later. Indeed, previous studies in midlife and older adults have observed chronotype-related cortical thinning in prefrontal areas, suggesting that cerebellar and ACC changes may serve as early neural markers of future prefrontal vulnerability.

Our findings contrast with other studies that reported reduced gray matter in posterior cortical regions among early chronotypes [8]. This discrepancy may stem from methodological differences, including the use of categorical vs. continuous chronotype classification, participant demographics (e.g., sex distribution), and neuroimaging pipelines. Nonetheless, both studies highlight the occipital cortex as a chronotype-sensitive region, reinforcing its relevance in the study of circadian neurobiology [7, 8]. Taken together, our results support the hypothesis that chronotype is associated with discernible neuroanatomical variation, even in the absence of clinical symptoms. These differences may underlie previously observed disparities in emotion regulation, attentional functioning, and cognitive performance between EC and LC individuals. Importantly, they suggest that sleep timing, beyond sleep duration or subjective quality, constitutes a critical dimension of brain health in young adulthood.

### Limitations

Several limitations should be considered when interpreting the current findings. First, although our sample size was adequate to detect regional structural differences, the generalizability of our results may be limited by the use of an open-access dataset with restricted diversity in terms of race, ethnicity, and socioeconomic background. Second, chronotype classification relied on self-report questionnaires, which, despite being validated, may not fully capture biological circadian markers. Third, our analyses focused exclusively on structural MRI data; integrating functional and diffusion-based imaging could provide a more comprehensive understanding of how chronotype affects brain connectivity and microstructure. Additionally, lifestyle-related variables such as body mass index, physical activity, caffeine or nicotine use were not available in the dataset, which limits our ability to account for other known modulators of brain morphology. Finally, while we employed a validated brain age prediction model, more granular and longitudinal approaches may be necessary to detect subtle deviations in aging trajectories and their relationship with chronotype.

## 5. Conclusion

This study provides new evidence that chronotype is associated with specific neuroanatomical differences in healthy young adults. Individuals with a late chronotype exhibited reduced gray matter volume in cerebellar regions and cortical thinning in the anterior cingulate and lateral occipital cortices—areas involved in cognitive control, emotion regulation, and sensory integration. These differences emerged in the absence of subjective sleep complaints or global brain aging, suggesting that circadian preference may shape brain structure independently of perceived sleep quality or duration. While no significant differences in estimated brain age were found, the regional morphological alterations observed in LC individuals may represent early neurobiological signatures of circadian misalignment. These findings highlight the potential value of sleep timing as a modifiable factor in neurodevelopmental and mental health trajectories. Future studies should employ multimodal and longitudinal designs to examine the stability, reversibility, and functional consequences of these structural patterns, and to determine whether targeted interventions aimed at improving circadian alignment can mitigate their impact.

## Institutional Review Board Statement

Not applicable.

## Data Availability Statement

The dataset analyzed in this study was obtained from the OpenNeuro repository (https://openneuro.org/datasets/ds003826/versions/1.1.0/3.0.0; accessed on March 1, 2025).

## References

1. Zou, H., et al., Chronotype, circadian rhythm, and psychiatric disorders: Recent evidence and potential mechanisms. Front Neurosci, 2022. 16: p. 811771.

2. Zheng, N.S., et al., Sleep patterns and risk of chronic disease as measured by long-term monitoring with commercial wearable devices in the All of Us Research Program. Nat Med, 2024. 30(9): p. 2648–2656.

3. Montaruli, A., et al., Biological Rhythm and Chronotype: New Perspectives in Health. Biomolecules, 2021. 11(4).

4. Gorfine, T., et al., Sleep-anticipating effects of melatonin in the human brain. Neuroimage, 2006. 31(1): p. 410–8.

5. Horne, C.M. and R. Norbury, Altered resting-state connectivity within default mode network associated with late chronotype. J Psychiatr Res, 2018. 102: p. 223–229.

6. Horne, C.M. and R. Norbury, Exploring the effect of chronotype on hippocampal volume and shape: A combined approach. Chronobiol Int, 2018. 35(7): p. 1027–1033.

7. Evans, S.L., et al., Evening preference correlates with regional brain volumes in the anterior occipital lobe. Chronobiol Int, 2021. 38(8): p. 1135–1142.

8. Rosenberg, J., et al., Chronotype differences in cortical thickness: grey matter reflects when you go to bed. Brain Struct Funct, 2018. 223(7): p. 3411–3421.

9. Grill-Spector, K., Z. Kourtzi, and N. Kanwisher, The lateral occipital complex and its role in object recognition. Vision Res, 2001. 41(10-11): p. 1409–22.

10. Malach, R., et al., Object-related activity revealed by functional magnetic resonance imaging in human occipital cortex. Proc Natl Acad Sci U S A, 1995. 92(18): p. 8135–9.

11. Lucas, R.J., et al., Measuring and using light in the melanopsin age. Trends Neurosci, 2014. 37(1): p. 1–9.

12. Schmahmann, J.D., The cerebellum and cognition. Neuroscience letters, 2019. 688: p. 62–75.

13. Stoodley, C.J. and J.D. Schmahmann, Functional topography in the human cerebellum: a meta-analysis of neuroimaging studies. Neuroimage, 2009. 44(2): p. 489–501.

14. Mishra, S., I. Beheshti, and P. Khanna, A review of neuroimaging-driven brain age estimation for identification of brain disorders and health conditions. IEEE Reviews in Biomedical Engineering, 2021. 16: p. 371–385.

15. Sone, D. and I. Beheshti, Neuroimaging-based brain age estimation: A promising personalized biomarker in neuropsychiatry. Journal of Personalized Medicine, 2022. 12(11): p. 1850.

16. Oginska, H., J. Mojsa-Kaja, and O. Mairesse, Chronotype description: In search of a solid subjective amplitude scale. Chronobiol Int, 2017. 34(10): p. 1388–1400.

17. Ogińska, H., Can you feel the rhythm? A short questionnaire to describe two dimensions of chronotype. Personality and Individual Differences, 2011. 50(7): p. 1039–1043.

18. Zareba, M.R., et al., Neuroimaging of chronotype, sleep quality and daytime sleepiness: Structural T1-weighted magnetic resonance brain imaging data from 136 young adults. Data Brief, 2022. 41: p. 107956.

19. Farokhian, F., et al., Comparing CAT12 and VBM8 for detecting brain morphological abnormalities in temporal lobe epilepsy. Frontiers in neurology, 2017. 8: p. 428.

20. Klein, A. and J. Tourville, 101 labeled brain images and a consistent human cortical labeling protocol. Front Neurosci, 2012. 6: p. 171.

21. Beheshti, I., S. Booth, and J.H. Ko, Differences in brain aging between sexes in Parkinson’s disease. npj Parkinson’s Disease, 2024. 10(1): p. 35.

22. Botvinick, M.M., J.D. Cohen, and C.S. Carter, Conflict monitoring and anterior cingulate cortex: an update. Trends Cogn Sci, 2004. 8(12): p. 539–46.

23. Bush, G., P. Luu, and M.I. Posner, Cognitive and emotional influences in anterior cingulate cortex. Trends Cogn Sci, 2000. 4(6): p. 215–222.

